# Factors Associated with Food Insecurity among Pregnant Women in Gedeo Zone Public Hospitals, Southern Ethiopia

**DOI:** 10.1101/2022.02.16.22271073

**Authors:** Abriham Shiferaw Areba, Arega Haile, Belayneh Genoro Abire, Berhanu Gidisa Debela, Miheret Tesfu Legesse, Sewitemariam Desalegn Andarge, Girum Gebremeskel Kanno, Belay Negassa Gondol, Desta Erkalo Abame

**Affiliations:** School of Public Health, College of Medicine and Health Science, Wachemo University, Hossana, Ethiopia; Department of Statistics, College of Natural Science, Dilla University, Dilla, Ethiopia; School of Public Health, College of Medicine and Health Science, Dilla University, Dilla, Ethiopia

**Author notes:** Corresponding Author: Abriham Shiferaw Areba.

**Keywords:** Food insecurity, Pregnant, Gedeo Zone, Public Hospital, Ethiopia

## Abstract

**Background:** Food insecurity refers to a lack of consistent access to sufficient food for active, better health. Around two billion people worldwide suffer from food insecurity and hidden hunger. Food insecurity and associated factors among pregnant women in Gedeo Zone Public Hospitals, Southern Ethiopia, are the focus of this study.

**Method:** From May to June 2021 G.C. institutional-based cross-sectional study was conducted among pregnant women in Gedeo zone public hospitals. A sample of 506 women has been used, and a multistage cluster sampling technique was used. An adjusted odds ratio (AOR) and their 95% confidence intervals (CI) were calculated to determine the association between various factors and outcomes. A p-value of less than 0.05 was considered significant in multivariable regression.

**Results:** Food insecurity was found to be prevalent in 67.4% of pregnant mothers. The results of a multivariable logistic regression revealed that pregnant women from rural areas [AOR=0.532, 95 % CI: 0.285, 0.994], married [AOR=0.232, 95% CI: 0.072, 0.750], have a secondary education [AOR=0.356, 95%CI: 0.154, 0.822], and be employed [AOR=0.453, 95% CI: 0.236, 0.872], income between 1000 and 2000 [AOR=0.163, 95% CI: 0.066, 0.399], and income greater than 2000 [AOR=0.125, 95% CI: 0.053,0.293], the wealth index middle and rich [AOR=0.441, 95% CI: 0.246, 0.793] were significant predictors of food insecurity among pregnant mothers [AOR=0.24, 95 % CI: 0.128, 0.449].

**Conclusion:** The study area had a high prevalence of food insecurity. Food insecurity was reduced in those who lived in rural areas, were married, had a secondary education, earned between 1000 and 2000 ETB and more than 2000 ETB, and had a wealth index of middle and rich.

## Introduction

Food insecurity refers to a lack of sufficient food, as well as restrictions on the quality, quantity, and/ or frequency of food consumption (1–4). Goal 2 of the Sustainable Development Goals (SDGs) aims to end hunger, increase food security, and promote sustainable agriculture. Target 2.1 of the SDGs is aimed at achieving the objective of ending hunger and ensuring year-round access to food for all people, including pregnant and lactating mothers, by 2030 (5).

At the worldwide level, gender differences in the incidence of moderate and severe food insecurity have expanded in the year of the COVID-19 pandemic, with women experiencing 10% more moderate or severe food insecurity in 2020 than males, compared to 6% in 2019. Severe food insecurity affects 28.7% of the population in Eastern Africa, while moderate to severe food insecurity affects 65.3 percent of the population (6).

According to the FAO estimate for 2021, over 2 billion people worldwide are afflicted by moderate levels of food insecurity and hunger, with Sub-Saharan Africa having the highest prevalence (21 percent of the population) (6,7). According to recent studies in Ethiopia, 7% of households experience severe food insecurity, while 11% and 22% of households experience mild and moderate food insecurity, respectively (8). Food insecurity is frequently seen as the most significant impediment to individual nutrition; as well as its effect on other health and behavior outcomes is increasingly being recognized (9–11). It can be understood and addressed as its entity but it should also be recognized as an important associate of health and nutrition outcomes (12, 13). In addition, to negative effects on mothers’ health, Food insecurity in pregnant women also has negative consequences for the child. According to various studies, Food insecurity has been linked to poor pregnancy outcomes, including low birth weight, gestational diabetes, and pregnancy problems (14-16). A study conducted in the United States showed that maternal food insecurity was associated with an increased risk of certain birth defects, such as cleft palate, transposition of the great arteries, and Anencephaly (17).

Furthermore, young children from food-insecure families have poorer general health (18–20), are more likely to be hospitalized (19, 21), have lower levels of parent-child attachment, and experience developmental delays (22–24). Food insecurity and food shortages are associated with poor general, mental and physical health in women. A study in the USA indicated that food insecurity was associated with women’s reduced mental health. Mental symptoms including depression, stress, and anxiety were associated with the household food insecurity in a dose-response relationship and were increased with worsening the food insecurity status (25).

Food insecurity has a substantial effect on the physical health of both pregnant women and their children, directly compromising the nutritional state and serum profile of micronutrients, such as iron. It may also trigger a series of stressful events in the family environment due to the difficulty in obtaining food, provoking deterioration in maternal mental health and consequent development of anxiety and depression, and also leading to negative outcomes concerning childcare (26).

Household food insecurity is expected to vary depending on the household head’s gender, age, and level of education; the size of the household; the quantity of livestock held; and financial and human capital-related issues (27). Because food security affects a pregnant woman’s nutritional condition, which is a significant environmental risk factor for poor pregnancy outcomes, securing a sufficient food supply for pregnant women has been a top priority for concerned parties. Although there have been studies in Ethiopia that focus on household food insecurity, it is critical to analyze the prevalence of pregnant food insecurity in the context of COVID 19 and its associated factors in the research region that has not been studied. As a result, the goal of his research was to support pregnant women who were food insecure during prenatal and postnatal care.

## Methods

### Study Design and Study Area Description

An institution-based cross-sectional study was conducted in Gedeo Zone Public Hospitals Southern Ethiopia. Gedeo zone is located 369 km from Addis Ababa to south on Addis Ababa-Moyale international road and 90 km from Hawassa (Capital city of the region) in south Nation Nationality and people regional state. The Zone has 1 referral hospital, 3 primary hospitals (Bule, Gedeb, and Yerga Chefe), 38 health centers, 146 health posts, 4 NGO clinics, and 17 reported private health facilities.

### Source Population and Study Population

All pregnant mothers attending antenatal care in Gedeo Zone Health facilities were the source population, while pregnant mothers attending antenatal care in Gedeo Zone Public Hospitals were the study population.

### Determination of Sampling Size and Sampling Procedure

The sample size used for this investigation was estimated and computed using a single population proportions method with the following assumptions: 324 percent FI among nursing moms, 95 percent confidence level of 1.96, the margin of error of 0.05, and design effect of 1.5. As a result, the study’s ultimate sample size was 506 participants (28). In terms of sampling, multistage cluster sampling entailed picking a sample in several steps. Pregnant mothers were the study’s subject. The pregnant women were sampled using a proportional sampling technique. After consent, one mother was randomly selected from among the pregnant women who matched the eligibility criteria to participate in the study. This approach was repeated until each woreda provided the required number of responders.

### Data collection and measurements

The study data collection instruments were developed after searching PubMed, Google Scholar, Hinari, and the Lancet series for various types of literature. The data was collected using a standardized interviewer-administered questionnaire. The questionnaire was written in English, then translated into Amharic, and then returned to English by language experts to ensure consistency and correctness. Six diploma nurses who were proficient in the local language (Gedeo’ufa) as data collectors and two BSc midwives as supervisors were hired based on their past data collecting experience.

Nine standard HFIAS questions derived from the FANTA project were used to determine the outcome variable food insecurity. The instrument consists of nine questions that illustrate the frequency of occurrence and quantify the severity of food insecurity in the previous four weeks using Likert Scale responses (0=Never, 1=rarely (1 or 2 times), 2=occasionally (3–10 times), and 3=frequently (>10 times). The pregnant ladies were required to respond to these questions on behalf of their entire household. At the time of data collection, this technique was used to assess food access for all family members. The nine items ranging from 0 to 27 were used to compute the cumulative score of food insecurity among expectant mothers, with a higher score indicating that the household members experienced more food insecurity. All ‘Yes’ responses were coded as ‘1’ and ‘No’ responses were coded as ‘0,’ and the responses were totaled to determine the level of household food insecurity (29).

The household’s wealth index was calculated using Principal Component Analysis (PCA) and took into account the latrine, water source, household assets, livestock, and ownership of agricultural land. All non-dummy variables’ responses were divided into three groups. The highest score was given a 1 rating. The two lower values, on the other hand, were given code 0. The variables with a commonality value larger than 0.5 were used to generate factor scores in PCA. Finally, the wealth score was calculated using each household’s score on the first major component. The wealth score was divided into three quintiles to classify households as low, medium, or wealthy.

### The Study’s Variables

The following are the response and predictor variables considered in the model for parameter estimation.

### Variable of Response

Food insecurity amongst pregnant women is the study’s outcome variable. If the women are food insecure, this can be dichotomized as 1 and 0 correspondingly.

The authors used a previous study conducted in California to assess food insecurity status, which is based on American standard food insecurity determination (30).

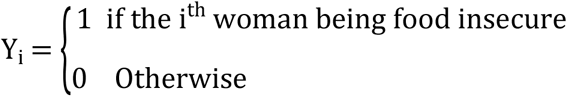

### Explanatory Variables

The table below lists the predictor variables that were investigated in this study to investigate food insecurity among pregnant women.

### Eligibility Criteria

#### Inclusion Criteria

All Pregnant mothers of pregnancy attending ANC service at selected health institutions were included in this study.

#### Exclusion Criteria

Pregnant mothers’ co-morbidities with complications and special requirements were excluded from the study. Diagnosed with chronic diseases like diabetes, hypertension, and twin pregnancy

### Operational Definitions

#### Food secure women

women who have experienced none of the FI (access) conditions or have just been worried, although rarely, during the past 4 weeks.

#### Food insecure women

women who are unable at all times to access food sufficient to lead an active and healthy life (includes all stages of FI; mild, moderate, and severe).

#### Mildly food insecure women

women who worry about not having enough food sometimes or often and/or are unable to eat preferred foods and/or eat a more monotonous diet than desired and/ or some foods considered undesirable, but only rarely.

#### Moderately food insecure women

women who sacrifice quality more frequently, by eating a monotonous diet or undesirable foods sometimes or often, and/or have started to cut back on quantity by reducing the size of meals or number of meals, rarely or sometimes. However, they do not experience any of the three most severe conditions.

#### Severely food insecure women

Women who have been forced to cut back on the meal size or number of meals often and/or experience any of the three most severe conditions (running out of food, going to bed hungry, or going a whole day and night without eating)

#### Wealth status

reliability test was performed using the economic variables involved in measuring the wealth. The variables which were employed to compute the principal component analysis, at the end of the principal component analysis, the wealth index was obtained as a continuous scale of relative wealth. Finally, Percentile groups of the wealth index were created to group under three wealth categories, poor, middle, and rich.

### Data quality control

Before data collection, the questionnaire was first written in English, then translated into Amharic, and then back to English for consistency. The purpose, methodology of the research on food insecurity, data collecting and interviewing style, and data recording were all covered in two days of training one week previous to the day of data collection. In a health center outside of the study area, the questionnaire was pre-tested on 5% of actual respondents. The supervisors and primary investigators kept a close eye on the overall activities during the data collection period to guarantee that the data was of high quality. Before analysis, all of the obtained data were double-checked, coded, entered into SPSS version 25, and cleaned to eliminate inconsistencies and incompleteness. The STATA/SE statistical software package version 14.0 was used to analyze the data.

### Data Analysis

Descriptive statistics were used to report the distribution of the data among variables using frequency and percentage. A bi-variable logistic regression analysis was performed to assess associations between each independent variable and food insecurity. A multivariable model should include all covariates that were relevant in bi-variable analyses at the P = 0.20 to 0.25 level from the start. In a multivariable model, the variables that tend to be relevant from the bi-variable analysis are fitted together. For multivariate analysis, statistical significance was determined using a 95 % confidence interval and a P-value of less than 0.05. As a consequence, the backward exclusion is used to omit non-significant variables from the final model (31).

## Results

A total of 506 pregnant moms were considered in this investigation. Food insecurity and food security were found in 67.4% and 32.6 percent of those moms, respectively. There were 139 (27.47 percent) and 367 (72.53 percent) women from rural and urban areas, respectively, with rural residents experiencing 108 (21.34 percent) less food insecurity than urban residents experiencing 233 (46.05 percent).

When it comes to the age of the mothers, the minimum number of women discovered in the age group of 15-19 years is 26 (5.14%), while the greatest number of women found in the age group of 20-24 years is 224. (44.27 percent). 478 (94.47 percent) of the moms in the research were married, whereas 28 (5.53) were unmarried (single and divorced) (Table 2).

**Table 1:** Description of independent variables used in the study

| Covariates | Description | Categories |
| --- | --- | --- |
| Age | Age of pregnant women | 0 = 15-19, 1 = 20-24, 2 = 25-29, 3= $\geq 30$ |
| Woreda | Woreda of pregnant women | 0= Dilla, 1= Gedebe, 2= Yirga Chafe, 3= Bule, 4= Others |
| Residence | Residence of women live | 0 = Urban 1=Rural |
| Education | Educational level of women | 0 = No education, 1= Primary, 2= Secondary, 3= Higher |
| Marital status | Marital status of women | 0= Others (Single, Divorce), 1= Married |
| Employment | Employment status of women | 0= Unemployed, 1= Employed |
| Income | Average Monthly Income | 1= < 1000, 2= 1000-2000, 3= >2000 |
| Wealth Index | Wealth Index of women | 1= Poor, 2= Middle, 3= Rich |
| Family Size | No. of family member | 1= 2, 2= 3-4, 3= 5 and Above |
| Parity | No. of live birth | 0= No Birth, 1= One Birth, 2= Two Birth, 3= three and above birth |
| Gravid | No. of pregnancy | 1= First pregnancy, 2= 2-3 Pregnancy, 3= 4 and above |
| Willingness | Pregnancy willingness | 0= Unwanted, 1= Wanted |
| Complication | Pregnancy Complication | 0= No, 1= Yes |
| Food Extension | Food Extension Service | 0= No, 1= Yes |

**Table 2:** Descriptive Summaries of Food Insecurity among Pregnant Mothers in Gedeo Zone Public Hospitals

| Covariates | Categories | Food Insecurity status |  | No. Out of 506 (%) |
| --- | --- | --- | --- | --- |
|  |  | Food Secure (%) | Food Insecure (%) |  |
| Residence | Rural | 31(6.13) | 108(21.34) | 139(27.47) |
|  | Urban | 134(26.48) | 233(46.05) | 367(72.53) |
| Age (year) | 15-19 | 8(1.58) | 18(3.56) | 26(5.14) |
|  | 20-24 | 75(14.82) | 149(29.45) | 224(44.27) |
|  | 25-29 | 61(12.06) | 106(20.95) | 167(33) |
|  | 30 and Above | 21(4.15) | 68(13.44) | 89(17.59) |
| Marital Status | Unmarried | 4(0.79) | 24(4.74) | 28(5.53) |
|  | Married | 161(31.82) | 317(62.65) | 478(94.47) |
| Educational status | No Education | 13(2.57) | 82(16.21) | 95(18.77) |
|  | Primary | 54(10.67) | 141(27.87) | 195(38.54) |
|  | Secondary | 55(10.87) | 77(15.22) | 132(26.09) |
|  | Higher | 43(8.50) | 41(8.10) | 84(16.60) |
| Employment status | Unemployed | 112(22.13) | 303(59.88) | 415(82.02) |
|  | Employed | 53(10.47) | 38(7.51) | 91(17.98) |
| Monthly Income | Less than 1000 | 7(1.38) | 104(20.55) | 111(21.94) |
|  | 1000-2000 | 35(6.92) | 84(16.60) | 119(23.52) |
|  | Greater than 2000 | 123(24.31) | 153(30.24) | 276(54.55) |
| Wealth Index | Poor | 24(4.74) | 144(28.46) | 168(33.20) |
|  | Middle | 58(11.46) | 111(21.94) | 169(33.40) |
|  | Rich | 83(16.40) | 86(17) | 169(33.40) |
| Family Size | 2 | 37(7.31) | 86(17) | 123(24.31) |
|  | 3-4 | 83(16.40) | 146(28.85) | 229(45.26) |
|  | 5 and Above | 45(8.89) | 109(21.54) | 154(30.43) |
| Parity | No Previous Birth | 50(9.88) | 112(22.13) | 162(32.02) |
|  | 1 Previous Birth | 54(10.67) | 86(17) | 140(27.67) |
|  | 2 Previous Birth | 29(5.73) | 59(11.66) | 88(17.39) |
|  | 3 and Above | 32(6.32) | 84(16.60) | 116(22.92) |
| Gravid | 1 Pregnancy | 45(8.89) | 110(21.74) | 155(30.63) |
|  | 2-3 Pregnancies | 87(17.19) | 147(29.05) | 234(46.25) |
|  | 4 and Above | 33(6.52) | 84(16.60) | 117(23.12) |
| Pregnancy willingness | Unwanted | 10(1.98) | 25(4.94) | 35(6.92) |
|  | Wanted | 155(30.63) | 316(62.45) | 471(93.08) |
| Complication of Pregnancy | No | 152(30.04) | 304(60.08) | 456(90.12) |
|  | Yes | 13(2.57) | 37(7.31) | 50(9.88) |
| Food Extension Service | No | 142(28.06) | 284(56.13) | 426(84.19) |
|  | Yes | 23(4.55) | 57(11.26) | 80(15.81) |
| Unmarried (Single, Divorced) |  |  |  |  |

A single covariate binary logistic regression model analysis is an appropriate approach for screening out potentially essential variables before including them directly in a multivariable model. Each covariate’s association to food insecurity among pregnant women was discussed. Food insecurity among pregnant mothers was significantly associated with place of residence, marital status, educational status, employment status, average monthly income, wealth index, family size, parity, and gravidity, but the age of mother, pregnancy willingness, pregnancy complications, and food extension service were not significant at a modest level of significance of 0.25.

In the Gedeo zone Public Hospitals, the place of residence, marital status, educational status, employment status, average monthly income, and wealth index all had statistically significant effects on food insecurity, i.e. the confidence interval for the adjusted odds ratio did not include one and the P-value was less than 0.05. The estimated odds ratios for pregnant mothers from rural areas were 0.532 when other predictor factors were kept constant in a multivariable regression model. This means that pregnant mothers from rural areas were 0.532 times (AOR=0.532, 95% CI: 0.285, 0.994) less likely than mothers from urban areas to be food insecure.

Food insecurity was reduced by 76.8% in expectant pregnant mothers who were married compared to those who were not (single, divorced) (AOR=0.232, 95 percent CI: 0.072, 0.750). When the influence of other factors was held constant in the model, pregnant mothers with secondary education had a 35 percent lower risk of food insecurity than pregnant mothers without a secondary education (AOR=0.35, 95 CI: 0.154, 0.822). With the effect of other independent variables constant in the model, pregnant mothers who had employment status were 0.453 times (AOR=0.453, 95% CI: 0.236 to 0.872) less likely to have food insecurity than those who had unemployment status.

When comparing women with an average monthly income of 1000 ETB to 2000 ETB and greater than 2000 ETB to women with an average monthly income of less than 1000 ETB, extreme food insecurity decreased by 16.3% and 12.3%, respectively (AOR=0.163, 95 percent CI: 0.066, 0.399 and AOR=0.123, 95 percent CI: 0.053, 0.293). When other predictor variables in the regression model were held constant, pregnant women with middle and high economic status were 0.441 and 0.24 times less likely to have food insecurity than those with low economic status (AOR=0.441, 95 percent CI 0.246 to 0.793) and (AOR=0.24, 95 percent CI 0.128 to 0.449) respectively (Table 3)

**Table 3:**
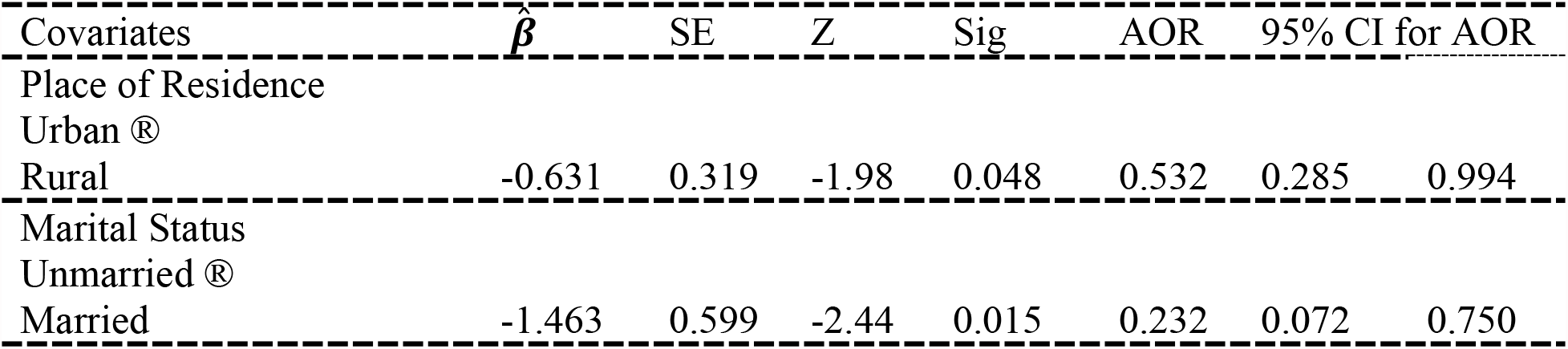
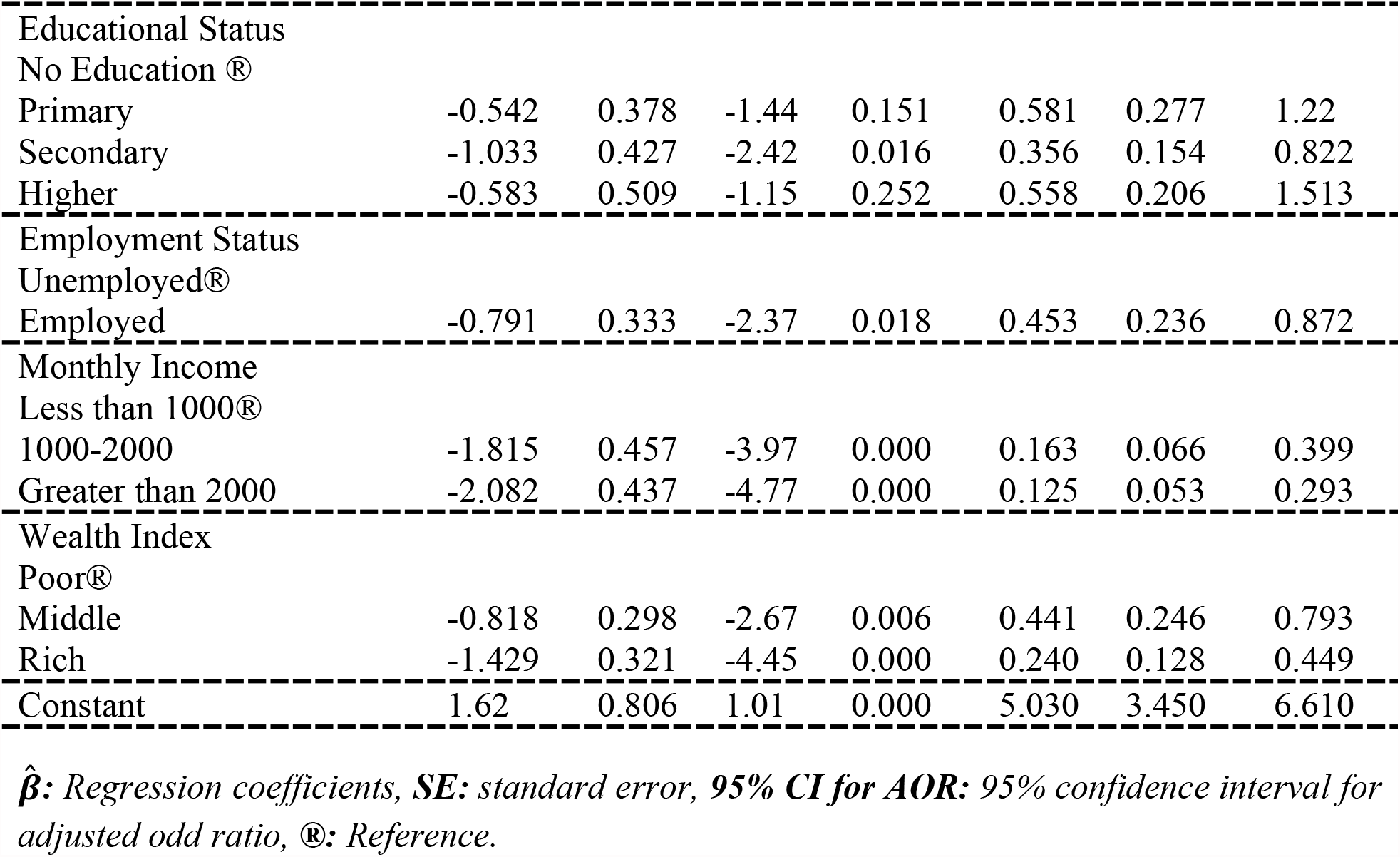
Multivariable Logistic Regression Analysis for Food Insecurity among Pregnant Mothers in Gedeo Zone Public Hospitals

| Covariates | $\hat{\beta}$ | SE | Z | Sig | AOR | 95% CI for AOR | |
| --- | --- | --- | --- | --- | --- | --- | --- |
| Place of Residence |  |  |  |  |  |  |  |
| Urban ® |  |  |  |  |  |  |  |
| Rural | -0.631 | 0.319 | -1.98 | 0.048 | 0.532 | 0.285 | 0.994 |
| Marital Status |  |  |  |  |  |  |  |
| Unmarried ® |  |  |  |  |  |  |  |
| Married | -1.463 | 0.599 | -2.44 | 0.015 | 0.232 | 0.072 | 0.750 |
| ----- |  |  |  |  |  |  |  |
| Educational Status |  |  |  |  |  |  |  |
| No Education® |  |  |  |  |  |  |  |
| Primary | -0.542 | 0.378 | -1.44 | 0.151 | 0.581 | 0.277 | 1.22 |
| Secondary | -1.033 | 0.427 | -2.42 | 0.016 | 0.356 | 0.154 | 0.822 |
| Higher | -0.583 | 0.509 | -1.15 | 0.252 | 0.558 | 0.206 | 1.513 |
| ----- |  |  |  |  |  |  |  |
| Employment Status |  |  |  |  |  |  |  |
| Unemployed® |  |  |  |  |  |  |  |
| Employed | -0.791 | 0.333 | -2.37 | 0.018 | 0.453 | 0.236 | 0.872 |
| ----- |  |  |  |  |  |  |  |
| Monthly Income |  |  |  |  |  |  |  |
| Less than 1000® |  |  |  |  |  |  |  |
| 1000-2000 | -1.815 | 0.457 | -3.97 | 0.000 | 0.163 | 0.066 | 0.399 |
| Greater than 2000 | -2.082 | 0.437 | -4.77 | 0.000 | 0.125 | 0.053 | 0.293 |
| ----- |  |  |  |  |  |  |  |
| Wealth Index |  |  |  |  |  |  |  |
| Poor® |  |  |  |  |  |  |  |
| Middle | -0.818 | 0.298 | -2.67 | 0.006 | 0.441 | 0.246 | 0.793 |
| Rich | -1.429 | 0.321 | -4.45 | 0.000 | 0.240 | 0.128 | 0.449 |
| ----- |  |  |  |  |  |  |  |
| Constant | 1.62 | 0.806 | 1.01 | 0.000 | 5.030 | 3.450 | 6.610 |
| ----- |  |  |  |  |  |  |  |

***$\hat{\beta}$** : Regression coefficients, **SE**: standard error, **95% CI for AOR**: 95% confidence interval for adjusted odd ratio, ®: Reference.*

## Discussion

The goal of this study was to find out how often food insecurity is among pregnant women and what factors contribute to it. Food security and proper nutrition are essential for human growth and development, which necessitates access to enough, diverse, and high-quality food resources (25). In terms of food insecurity, 67.4 percent of pregnant women in this survey were food insecure. The findings of this study were similar to those of studies conducted in Hossana (67.5 percent) (32) and Areka (69.6 percent) (33). On the other hand, it is greater than the Atay District (36.8%) (34), Abay District (38.1%) (26), and Sodo town (37.6%) studies completed in Ethiopia (35).

Seasonal variations in family food security status, which are frequently higher in Ethiopia’s summarizing season, could explain the increased degree of food insecurity. It could also be explained by households having fewer and smaller meals as a result of a monotonous diet and a lack of variety in food items. These discrepancies could be related to variances in the study participants’ socio-demographic variables. Seasonal fluctuation may be another major explanation for the apparent difference, as this study was conducted during the summer season whereas the other experiments were conducted during the pre-harvest season.

According to the findings, rural residency, marriage, secondary education, average monthly income between 1000 and 2000 ETB and greater than 2000 ETB, and wealth index intermediate and rich were all significant predictors of food insecurity among pregnant women. The location of residence was found to be a major differential for food insecurity in this study, and the findings suggest that moms from urban areas are more likely to have food insecurity than mothers from rural areas. Previous research backs up this conclusion (36).

Pregnant mothers who were part of a married pair were less likely to be food insecure than those who were single or divorced. This could be because married households in the study area had better access to farmland and social security than unmarried households. This was supported by research (35, 37). Mothers’ educational status is one of the determinants of their food insecurity.

This implies that women with secondary education were less likely to be food insecure than those without. Previous research backs up this research (25, 34, 37, 38, 39). This is because educated mothers are more likely to know how to create, improve, manage, and produce enough varieties of farms to ensure their families and their food security. Employment status is one factor that influences food insecurity among pregnant women. The findings reveal that expectant moms who are employed are less likely to be food insecure than those who are unemployed. The findings of this study were similar to those of others (25, 32, 39, 40).

Pregnant women with an average monthly income of 1000 ETB to 2000 ETB and more than 2000 ETB experienced 16.3% and 12.3% reductions in food insecurity, respectively, when compared to pregnant women with an average monthly income of less than 1000 ETB. i.e., pregnant mothers with a higher average monthly income had much less food insecurity than those with a lower average monthly income (34, 38, 39). This is because women with a higher income can fight food insecurity by having a secondary source of food when there is food in the house.

Pregnant women with medium and upper-class economic positions were less likely to experience food insecurity than mothers with lower-class economic status. I.e. poor pregnant women were more likely to be food insecure as a result of their low wealth index. This could be explained by the fact that poor pregnant women may have no or only one source of income, making it difficult for them to purchase appropriate foods to meet the demands of their household members owing to extreme poverty. This finding was in line with earlier research (32, 33, 34, 41).

## Conclusion

From May to June, this study employed a food insecurity dataset collected from expectant women at Gedeo Zone Public Hospital. The study comprised 506 pregnant women, with 67.4% of them being food insecure and the remaining 36.6 percent being food secure. The results of a binary multivariable logistic regression model revealed that place of residence, marital status, educational status, employment status, average monthly income, and wealth index had statistically significant effects on food insecurity among pregnant mothers in Gedeo zone Public Hospitals. Rural residence, married, secondary education level, income level between 1000 ETB and 2000 ETB and greater than 2000 ETB, and wealth index intermediate & rich were reduce significant predictors of food insecurity.

## Data Availability

All relevant data are within the manuscript and its Supporting Information files

## Acknowledgments

The authors would like to express their gratitude to Dilla University for their financial support. We also like to express our gratitude to the data collectors and supervisors.

## Ethical Approval

Ethical approval of this study was approved after review by the “Dilla University Institutional Research and Ethical Review Board,” and the research was carried out only after an ethical letter was obtained. A consent form was provided to all participants, and the purpose and importance of the study were explained to each study participant. To ensure the confidentiality of the participant’s information, codes were used instead of the names of the participant. Participants were interviewed alone to maintain privacy. All participants did not pay for the test. Voluntary participation was clearly explained to all the participants that they could choose to participate in before the study.

## References

1. Blumberg SJ, Bialostosky K, Hamilton WL, Briefel RR. The effectiveness of a short form of the household food security scale. Am J Public Health. 1999; 89:1231–4.

2. Tarasuk VS. Household food insecurity with hunger is associated with women’s food intakes, health and household circumstances. J Nutr. 2001; 131:2670–6.

3. Bhattacharya J, Currie J, Haider S. Poverty, food insecurity, and nutritional outcomes in children and adults. J Health Econ. 2004; 23: 839–62.

4. Kaiser LL, Melgar-Quinonez H, Townsend MS, Nicholson Y, Fujii ML, Martin AC, Lamp CL. Food insecurity and food supplies in Latino households with young children. J Nutr Educ Behav. 2003; 35:148–53.

5. United Nations: Transforming our world: the 2030 Agenda for Sustainable Development. September 2015, New York

6. FAO, IFAD, UNICEF, WFP and WHO. 2021. The State of Food Security and Nutrition in the World 2021. Transforming food systems for food security, improved nutrition and affordable healthy diets for all. Rome, FAO. https://doi.org/10.4060/cb4474en

7. FAO, Ifad, UNICEF, W.P. & WHO. The State of Food Security and Nutrition in the World 2019. Safeguarding against economic slowdowns and downturns. CC, Rome, FAO. License.

8. Girma Gezimu Gebre and Dil Bahadur Rahut. Prevalence of household food insecurity in East Africa: Linking food access with climate vulnerability Climate Risk Management 33 (2021) 100333 Available online 4 June 2021

9. Core indicators of nutritional state for difficult to sample populations. J Nutr 1990; 120 (Sup l 11):1559S–600S.

10. FAO. The state of food insecurity in the world: addressing food insecurity in protracted crises. Rome, Italy: FAO, 2010.

11. Broca SS. Food insecurity, poverty and agriculture: a concept paper. Rome, Italy: FAO, 2002.

12. Pinstrup-Andersen P. Food security: definition and measurement. Food Security 2009;1:5– 7

13. Campbell CC. Food insecurity: a nutritional outcome or a predictor variable? J Nutr 1991; 121:408–15.

14. Borders AE, Grobman WA, Amsden LB, et al. Chronic stress and low birth weight neonates in a low-income population of women. Obstet Gynecol 2007; 109(2 Pt 1):331–8.

15. Laraia BA, Siega-Riz AM, Gundersen C. Household food insecurity is associated with self-reported pregravid weight status, gestational weight gain, and pregnancy complications. J Am Diet Assoc 2010;110:692–70

16. Laraia BA, Siega-Riz AM, Gundersen C. Household food insecurity is associated with self-reported pregravid weight status, gestational weight gain, and pregnancy complications. J Am Diet Assoc 2010;110: 692–701)

17. Carmichael SL, Yang W, Herring A, Abrams B, Shaw GM. Maternal food insecurity is associated with an increased risk of certain birth defects. J Nutr 2007; 137:2087–92.

18. Cook JT, Black M, Chilton M, et al. Are food insecurity’s health impacts underestimated in the U.S. population? Marginal food security also predicts adverse health outcomes in young U.S. children and mothers. Adv Nutr 2013; 4:51–61.

19. Cook JT, Frank DA, Levenson SM, et al. Child food insecurity increases risks posed by household food insecurity to young children’s health. J Nutr 2006; 136:1073–6.

20. Bronte-Tinkew J, Zaslow M, Capps R, et al. Food insecurity works through depression, parenting, and infant feeding to influence overweight and health in toddlers. J Nutr 2007; 137:2160–5.

21. Cook JT, Frank DA, Berkowitz C, et al. Food insecurity is associated with adverse health outcomes among human infants and toddlers. J Nutr 2004; 134:1432–8.

22. Zaslow M, Bronte-Tinkew J, Capps R, et al. Food security during infancy: implications for attachment and mental proficiency in toddlerhood. Matern Child Health J 2009; 13:66–80.

23. Hernandez DC, Jacknowitz A. Transient, but not persistent, adult food insecurity in?uences toddler development. J Nutr 2009; 139:1517–24.

24. Rose-Jacobs R, Black MM, Casey PH, et al. Household food insecurity: associations with at-risk infant and toddler development. Pediatrics 2008; 121:65–72.

25. Moafi, F. et al. ‘The relationship between food security and quality of life among pregnant women. 2018; 1–9.

26. Shone, M. et al. Household food insecurity and associated factors in West Abaya district. Agriculture & Food Security. 2017; 1–9. doi: 10.1186/s40066-016-0080-6.

27. Girma Gezimu Gebre and Dil Bahadur Rahut. Prevalence of household food insecurity in East Africa: Linking food access with climate vulnerability Climate Risk Management 33 (2021) 100333 Available online 4 June 2021

28. Nantale G, Tumwesigye NM, Kiwanuka N, Kajjura R. Prevalence and Factors Associated with Food Insecurity among Women Aged 18-49 Years in Kampala Slums Uganda ; A Mixed Methods Study. 2017;5(4):120–8.

29. Coates J, Bilinsky P, Coates J. Household Food Insecurity Access Scale (HFIAS) for Measurement of Food Access : Indicator Guide VERSION 3 Household Food Insecurity Access Scale (HFIAS) for Measurement of Food Access : Indicator Guide VERSION 3. 2007;(August).

30. Ghose et al. (2016), Association between food insecurity and anemia among women of reproductive age. PeerJ 4:e1945; DOI 10.7717/peerj.1945

31. Stevenson, M., 2009. An Introduction to Survival Analysis *., pp.1–32.

32. Asnakew Mekuria (2015) ‘Food Insecurity : Prevalence and Associated Factors among Adult Individuals Receiving Highly Active Antiretroviral Therapy (HAART) in ART Clinics of Hosanna Town. Open Access Library Journal Food. doi: 10.4236/oalib.1101800.

33. Habte Samuel, Gudina Egata, Wondimagegn Paulos, TesfahunYonas Bogale, M. T. (2019) ‘Food Insecurity and Associated Factors Among Households in Areka Town, Southern Ethiopia’, Journal of Health, Medicine and Nursing, 66, pp. 42–50. doi: 10.7176/JHMN.

34. Getacher, L. et al. (2020) ‘Food insecurity and its predictors among lactating mothers in North Shoa Zone, Central Ethiopia : a community sectional study based cross--’, pp. 1–9. doi: 10.1136/BMJopen-2020-040627.

35. Tadesse, A. ‘Household Food Insecurity and Associated Factors among Households in Sodo Town, 2015’, Food Science and Quality Managemen, 56, pp. 10–20.

36. Ukegbu, P. et al. (2019) ‘Food Insecurity and Associated Factors Among University Students’, 40(2), pp. 271–281. doi: 10.1177/0379572119826464.

37. Goldberg, S. L. and Mawn, B. E. (2020) ‘Predictors of Food Insecurity among Older Adults in the United States. doi: 10.1111/phn.12173

38. Abrahams, Z. et al. (2018) ‘Factors associated with household food insecurity and depression in pregnant South African women from a low socio-economic setting : a cross-sectional study, Social Psychiatry and Psychiatric Epidemiology, 53(4), pp. 363–372. doi: 10.1007/s00127-018-1497-y.

39. Dadras, A. O. et al. (2020) ‘The prevalence and associated factors of adverse pregnancy outcomes among Afghan women in Iran ; the possible impacts of domestic violence, poor mental health, housing issues, and food security.

40. Tantu, A. T., Gamebo, T. D. and Sheno, B. K. (2017) ‘Household food insecurity and associated factors among households in Wolaita Sodo town, 2015’, Agriculture & Food Security, pp. 1–8. doi: 10.1186/s40066-017-0098-4.

41. Shakiba, M. and Salari, A. (2021) ‘Food insecurity status and associated factors among rural households in the north of Iran’, (March). doi: 10.1177/0260106021996840.

